# Maternal immunisation against Group B Streptococcus: a global analysis of health impact and cost-effectiveness

**DOI:** 10.1101/2022.07.11.22277482

**Authors:** Simon R. Procter, Bronner P. Gonçalves, Proma Paul, Jaya Chandna, Farah Seedat, Artemis Koukounari, Raymond Hutubessy, Caroline Trotter, Joy E Lawn, Mark Jit

## Abstract

**Background:** Group B Streptococcus (GBS) can cause invasive disease (iGBS) in young infants, typically presenting as sepsis or meningitis, and is also associated with stillbirth and preterm birth. GBS vaccines are under development, but their potential health impact and cost-effectiveness have not been assessed globally.

**Methods:** We assessed the health impact and value (using net monetary benefit, NMB, which measures both health and economic effects of vaccination into monetary units), of GBS maternal vaccination across 183 countries in 2020. Our analysis uses a decision-tree model, combining risks of GBS-related outcomes from a Bayesian disease burden model with estimates of GBS related costs and Quality-Adjusted Life Years (QALYs) lost. We assumed 80% vaccine efficacy against iGBS and stillbirth, following the WHO Preferred Product Characteristics, and coverage based on the proportion of pregnant women receiving at least four antenatal visits. One dose was assumed to cost $50 in high-income countries, $15 in upper-middle income countries, and $3.50 in low-/lower-middle income countries. We estimated NMB using alternative normative assumptions that may be adopted by policy makers.

**Findings:** Vaccinating pregnant women could avert 214,000 (95% uncertainty range 151,000 – 457,000) infant iGBS cases, 31,000 deaths (14,000 – 67,000), 21,000 (9,000 – 52,000) cases of neurodevelopmental impairment, and 23,000 (10,000 – 58,000) stillbirths. A vaccine effective against GBS-associated prematurity might also avert 172,000 (13,000 – 378,000) preterm births. Globally, a 1-dose vaccine programme could cost $1.7 billion but save $385 million in healthcare costs. Estimated global NMB ranged from $1.1 billion ($-0.2 – 3.8 billion) to $17 billion ($9.1 – 31 billion).

**Interpretation:** Maternal GBS vaccination could have a large impact on infant morbidity and mortality globally and at reasonable prices is likely to be cost-effective.

## INTRODUCTION

*Streptococcus agalactiae*, commonly known as Group B Streptococcus (GBS), is an important bacterial pathogen causing morbidity and mortality in pregnant women and their babies and is also increasingly recognised as a cause of disease in non-pregnant adults.^1–3^ Invasive GBS (iGBS) disease in neonates and young infants can result from maternal colonisation and vertical transmission or environmental exposure after birth. It is classified by age at onset with early-onset GBS (EOGBS) occurring in the first 6 days of life, and late-onset disease (LOGBS) occurring between ages 7 and 89 days, and typically presents as sepsis, meningitis, or pneumonia. In 2020, an estimated 20 million pregnant women globally were colonised with GBS resulting in 231,000 (114,000 – 455,000) cases of EOGBS and a further 162,000 (70,000 – 394,000) LOGBS cases.^1^ Together these were estimated to have caused 58,000 to 91,000 infant deaths depending on the assumptions made about mortality in cases without access to healthcare. Furthermore, survivors of iGBS are at risk of long-term neurological sequelae with an estimated 37,100 (14,600 – 96,200) surviving infants developing moderate or severe neuro-developmental impairment (NDI).^1,4^ Maternal colonisation with GBS is also an important cause of adverse pregnancy outcomes with an estimated 46,000 (20,000 – 111,000) GBS stillbirths and is potentially linked with 518,000 (36,000 – 1,142,000) excess preterm births.

Currently the main strategies for preventing iGBS are based on intrapartum antibiotic prophylaxis (IAP). Many higher-income countries have reduced EOGBS incidence through IAP with eligible pregnant women identified either through risk-factor based screening or routine testing based on microbiological culture.^5^ Despite this success IAP has several limitations, notably it is not effective against LOGBS or GBS-associated stillbirths. In addition, the need for access to laboratory testing for microbiological screening based strategies, and the requirement to deliver antibiotics intravenously substantially limits the prospect of attaining high IAP coverage in many low-resource settings where the burden of iGBS is highest.^5^ There are also concerns that routine administration of antibiotics could contribute to antimicrobial resistance and might also have unintended impacts on the gut microbiota of newborns.^6^ Hence, there is substantial interest in alternative approaches to prevention.

Maternal immunisation is a potential alternative strategy whereby vertical transfer of antibodies in utero from a woman vaccinated during pregnancy affords protection to the mother, unborn foetus and newborn infant.^7^ Maternal immunisation with Tetanus Toxoid has been successfully used to reduce the burden of neonatal tetanus since the 1970s and, in the last decade, countries have been increasingly recommending routine vaccination of pregnant women against influenza and pertussis.^8^ In 2015 development of a maternal vaccine against GBS was identified as a priority by the WHO Product Development for Vaccines Advisory Committee (PDVAC),^9^ and three GBS maternal vaccine candidates have progressed to Phase-2 clinical trials.^10^ In 2021, the licensure of an affordable GBS vaccine by 2026 was identified as a key milestone in the WHO global roadmap for Defeating Meningitis by 2030.^11^

There have been previous economic evaluations of maternal GBS vaccination in the United States,^12–14^ Europe,^15–17^ and Sub-Saharan Africa.^18–20^ However, none of these studies have estimated the value of GBS vaccination in all world regions. A global economic evaluation of GBS vaccination is important to drive investment into vaccine development by indicating the vaccine’s potential value in different markets. It would also enable financing and pricing mechanisms to be put in place for equitable access to the vaccine once it is available. Such an evaluation is central to a Full Value of Vaccines Assessment (FVVA), which WHO has identified as key to catalysing vaccine development and subsequent equitable access.^21,22^ To inform the WHO GBS vaccine FVVA,^23^ we conducted the first global economic evaluation of maternal GBS vaccination in 183 countries, drawing on recently updated global disease burden estimates for GBS.^1^

## METHODS

### Model overview

We developed a decision-tree model (Fig. 1) to assess the health impact and cost-effectiveness of maternal vaccination against GBS in an annual cohort of pregnant women and their babies for the year 2020 compared with current practice of no vaccination. The size of the cohort of women in each country was calculated by combining country-specific estimates of the number of births from the United Nations World Populations Prospects (UNWPP)^24^ together with the stillbirth risk from the WHO Global Health Observatory.^25^ Our analysis included the 183 countries out of 195 UN member states for which UNWPP birth data was available, which excludes countries with estimated populations below 90,000.

**Figure 1:**
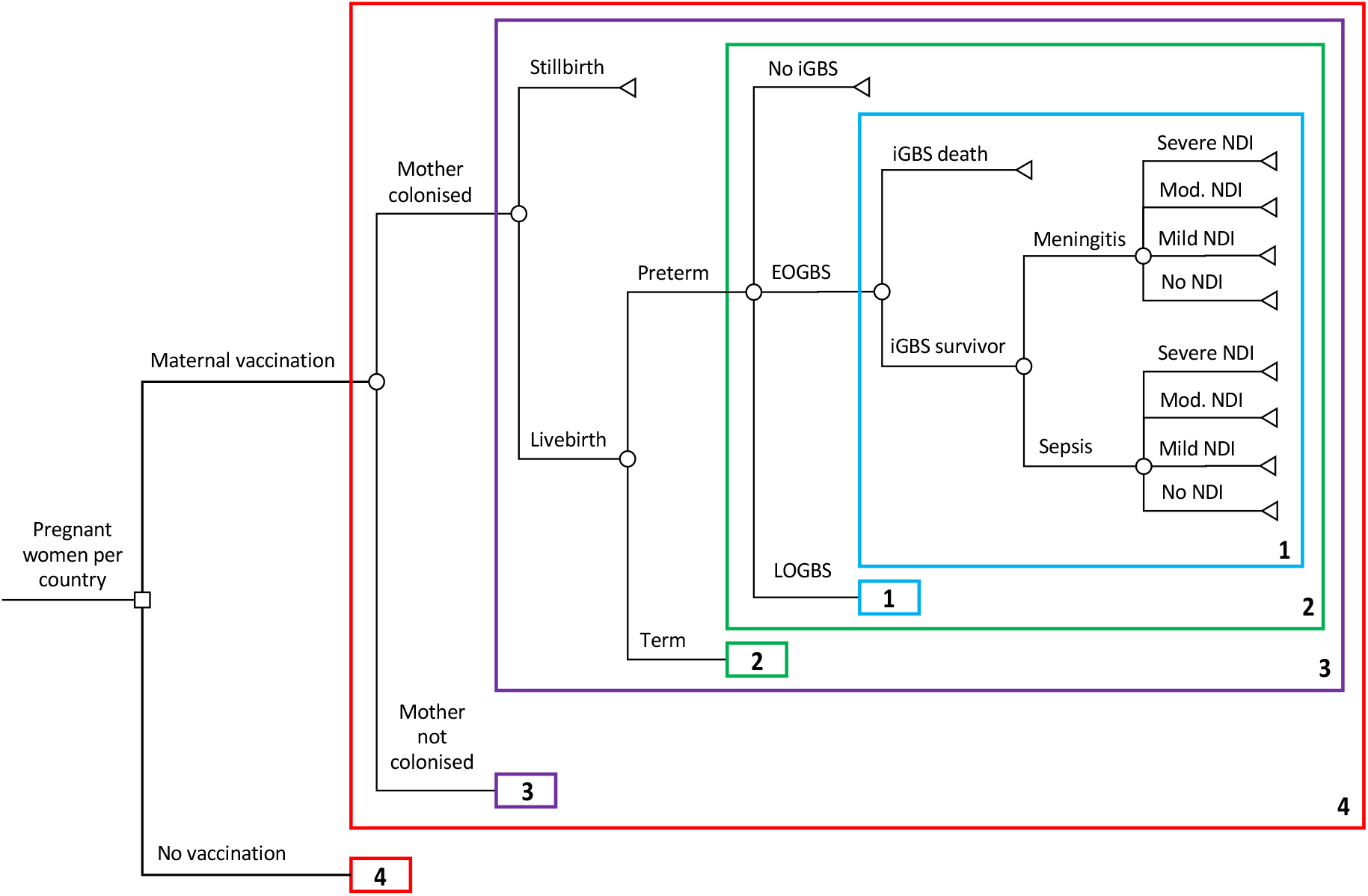
Decision tree for GBS-related outcomes in children for an annual birth cohort in 183 countries comparing maternal vaccination against no vaccination (current standard of care) Numbered boxes represent repeated model structure, however the risks for some outcomes vary across repeated branches.

The health impact model structure was designed to reflect the natural history of pregnancy related GBS infections and was aligned with the modelling framework used in recently reported global estimates of GBS burden.^1,26^ The model first stratifies pregnant women based on GBS colonisation status, and then by whether pregnancy results in a live birth or stillbirth. Live births are further sub-divided into preterm and term births, with infants then at risk of developing either EOGBS or LOGBS; the risk of EOGBS amongst babies born to non-colonised mothers was assumed zero. Invasive GBS disease (EOGBS or LOGBS) may then result in death or, amongst survivors of GBS sepsis or meningitis, either full recovery or long-term neurodevelopmental impairment.

The analysis used the lifetime of babies as the analytical time-horizon with health costs and Quality Adjusted Life Years (QALYs) calculated over the lifetime of infant iGBS survivors using country-specific life expectancy at birth.^24^ The model was used to compare scenarios with vaccination plus current practice against current practice without vaccination (i.e. assuming no change in each country’s IAP policy following vaccine introduction). All analyses were performed using R version 4.0.2. A Consolidated Heath Economic Evaluation Reporting Standards (CHEERS) checklist is provided in supplementary appendix 1.

### Disease risk

Model inputs are summarised in supplementary appendix A2. We parameterised the probability of different GBS-related outcomes in our model using posterior samples of key epidemiological parameters from the global burden estimates reported by Gonçalves et al.^1^ We used country-specific estimates of the prevalence of maternal GBS colonisation and of the risk of EOGBS in infants born to colonised mothers. The risks of LOGBS were then calculated using region-specific estimates of the fraction of iGBS cases that are EOGBS vs LOGBS. Regional classifications were based on the United Nations (UN) Sustainable Development Goal (SDG) region definitions.^27^

Case fatality risks (CFR) for EOGBS and LOGBS were also based on regional level estimates from Gonçalves et al. There are no data on CFRs for infants with EOGBS without access to care, so the authors considered two scenarios where they had either 90% CFR (following the approach of Seale and co-workers^3^) or the same CFR as other infants with EOGBS. In our analysis, we assumed in the base case that these infants had the same CFR as other infants with EOGBS, to be conservative about this highly uncertain parameter and because mothers of these children might also be less likely to receive maternal vaccines.

Among iGBS survivors the proportion of sepsis and meningitis, and the excess risk of mild, moderate, and severe NDI outcomes after meningitis were based on pooled global estimates, while NDI risks after sepsis were based on separate estimates for high-income and for low- and middle-income countries. The excess risk attributable to iGBS exposure was calculated assuming a counterfactual risk of mild or moderate and severe NDI amongst unexposed children from a large Danish cohort study.^28^ We based the proportion of moderate and severe NDI that was severe on the same study. Following the approach used in the burden estimation, our base case analysis included only the excess risk of moderate or severe NDI, which is likely to be more consistent across settings, but include mild NDI as a sensitivity analysis.^1,4^

To estimate country-specific GBS-associated stillbirth risk, national stillbirth estimates from the WHO Global Health Observatory^25^ were combined with regional estimates of the proportion of stillbirths caused by GBS.^1^ For the risk of GBS-associated prematurity we used national data on the proportion of preterm births^29^ together with the global odds ratio for the association between GBS maternal colonisation and preterm births.^1^ Further details on these calculations are provided in supplementary appendix A2.3 and A2.4.

### Health Outcomes

To calculate QALYs we assumed country-specific life-expectancy at birth for both normal births and survivors of iGBS. For term births we assumed no reduction in Health-Related Quality-of-Life (HRQoL), but for preterm births we applied a utility decrement over the child’s lifetime based on a systematic review and meta-analysis by Petrou et al.^30^ For the acute iGBS episode we approximated QALY loss assuming 29 days duration based on the average length-of-stay among studies in a recent systematic review of the acute costs of infant sepsis and meningitis,^31^ and applied health state utility decrements for hospitalisation with acute sepsis or meningitis from a US study in young children.^32^ For survivors with long-term sequelae, we applied utility decrements for mild, moderate, and severe NDI to each year of life and conservatively, given previous studies provide evidence of post-acute mortality after bacterial meningitis,^33,34^ assumed no change in life-expectancy. These utility values were based on a UK study, which assessed HRQoL in a cohort of children with NDI followed up at age 11.^35^

### Vaccination

Although clinical studies have demonstrated immunogenicity of candidate GBS vaccines, to date there have been no phase-3 efficacy trials.^10^ We therefore based our assumptions about vaccine efficacy (VE) and other characteristics of a GBS vaccine on the WHO preferred product characteristics (PPCs).^36^ In our base case we assumed a single-dose vaccine with 80% efficacy against both infant iGBS disease and GBS-stillbirth across all GBS serotypes. We also assumed no effect on GBS-associated prematurity because (i) the WHO PPC does not specify that GBS vaccines must reduce colonisation, which is most likely pathway for preventing GBS-associated prematurity, and (ii) the association between GBS maternal colonisation and higher risk of prematurity may be confounded.^37^ It is likely that delivery of GBS vaccines will need to be timed in either the late second trimester or third trimester and could be delivered through existing routine antenatal care (ANC) services. Hence, we assumed vaccine coverage based on the proportion of pregnant women in each country who attend at least four ANC visits (ANC4).^25^

We also considered a range of alternative scenarios (supplementary table 4): higher vaccine coverage based on the proportion of women attending at least one ANC visit (ANC1); a two-dose regimen; lower and higher VE (60% and 90%); and a vaccine that is also effective against GBS-associated prematurity. For the latter scenario we estimated the proportion of preterm births that are potentially protected through vaccination by combining the distribution of preterm births by gestational age^38^ with the timing of vaccine visits based on country-specific ANC data^39^ (supplementary appendix A2.5).

### Costs

Our analysis was undertaken from a healthcare payer economic perspective, and all costs are reported in 2020 United States Dollars (USD). Where cost inputs were for different years they were inflated using the World Bank Gross Domestic Product (GDP) deflator.^40^ Costs reported in different currencies were then converted to 2020 USD using historical foreign exchange rates.^41^ To estimate acute healthcare costs, we combined one GBS-specific cost estimate from a study in the United Kingdom,^42^ with the findings from a systematic review on the acute costs of infant sepsis and meningitis^43^, and result of a recent study reporting the acute costs of neonatal bacterial sepsis and meningitis in Mozambique and South Africa.^44^ We used linear regression to extrapolate country-specific cost estimates using total per capita healthcare expenditure as a predictor (supplementary appendix A2.6.) For long-term costs, no direct GBS-specific estimates exist in the literature. nnual costs amongst survivors with moderate and severe NDI were parameterised as a fixed proportion of between 4% and 28% of the acute cost estimate in each country, based on the range between a UK study of costs in children with NDI^35^ and a US study of costs in adults with disabilities.^45^

For the vaccine programme costs we extrapolated results from a systematic review of maternal vaccination delivery costs using regression against GDP per capita (supplementary appendix A2.7).^31^ We used previously estimated vaccine prices by World Bank country income group, which were based on a combination of price benchmarking against other vaccines and cost of goods analysis: $50 for high-income countries; $15 for upper-middle-income countries; and $3.50 for lower-middle-income and low-income countries.^46^

### Normative assumptions

A health intervention may be considered cost-effective if the cost per QALY gained falls below that country’s cost-effectiveness threshold. Here we use two commonly cited thresholds: (i) country gross domestic product per capita,^47^ (ii) published thresholds based on empirical estimates of the health opportunity cost of health care spending (supplementary appendix A2.8).^48,49^

A second normative assumption is the QALY loss attributed to a stillbirth. In many settings these are not assigned any health or disability weight, but it has been argued that they should be assigned a QALY loss close or the same as that of the death of a newborn.^50^ Here we consider two scenarios, one in which stillbirths are not assigned any QALY loss, and a second in which they are assigned the same QALY loss as the death of a newborn.

Following WHO guidelines, we discount costs at 3% and health effects at both 0% and 3% in alternative scenarios.^51^ Table 1 summarises the normative scenarios used.

**Table 1:** Parameter values used for least and most favourable normative assumptions.

| Parameter | Least favourable Assumptions | Most favourable assumptions |
| --- | --- | --- |
| Discount rate | 3% for costs and benefits | 3% for costs; 0% for benefits |
| Inclusion of stillbirth quality-adjusted life-years | Not included | Included |
| Cost-effectiveness threshold | Based on empirical estimates* | 1 x GDP per capita |

\*Cost-effectiveness thresholds were based either on estimates from Ochalek et al. and Woods et al. See supplementary appendix A2.8 for more detail.

### Economic analysis

To assess the cost-effectiveness of GBS maternal vaccination compared to current practice, we follow a Net Monetary Benefit (NMB) approach in which both the health and fiscal benefits of vaccination are expressed in monetary units.^52^ To calculate the NMB the incremental benefits in QALYs are multiplied by a country-specific cost-effectiveness threshold (CET; either empirical or 1 x GDP per capita) in USD and then the incremental costs are subtracted. An intervention may be considered cost-effective if the NMB is positive, since this is mathematically equivalent to the incremental cost-effectiveness ratio (ICER) being less than the CET. In addition to NMB, we also estimated the threshold price per dose at which a GBS vaccine would be cost-effective in each country.

An advantage of adopting an NMB framework is that our estimates for individual countries can be directly combined to estimate the aggregate value of vaccination both regionally and globally. To account for parameter uncertainty, for each scenario we ran 4000 simulations per country and calculated the median and the 95% uncertainty range (UR) based on 2.5 and 97.5 percentiles of the simulations. At the country level we also calculated the probability maternal GBS vaccination was cost-effective (i.e., the proportion of simulations with NMB > 0).

### Role of the funding source

The funder of the study had no role in study design, data collection, data analysis, data interpretation, or writing of the report.

## RESULTS

We estimate that vaccinating 99.8 million pregnant women across 183 countries could cost $1.7 billion but could save around $300 million in acute healthcare costs and $85 million in long-term healthcare costs, although these estimates have wide uncertainty. Overall, the incremental cost of GBS vaccination is about $1.3 billion, with the biggest cost increase in Europe and Northern America (Table 2).

**Table 2:** Annual global and regional impact of GBS maternal vaccination compared with no vaccination for the year 2020. All values are reported to 3 significant figures. Values in brackets are 95% uncertainty ranges. GBS = Group B Streptococcus; EOGBS = Early-Onset GBS; LOGBS = Late-Onset GBS; NDI = Neurodevelopmental Impairment; QALY = Quality Adjust Life Year. ^*^in scenario analysis where vaccine is assumed to have 80% VE against GBS associated prematurity ^^^global median values do not exactly equal the sum of the regional median values

| Description | Central & Southern Asia | Eastern & South-Eastern Asia | Europe & Northern America | Latin America & Caribbean | Northern Africa & Western Asia | Oceania | Sub-Saharan Africa | Global<sup>a</sup> |
| --- | --- | --- | --- | --- | --- | --- | --- | --- |
| Number of women vaccinated (millions) | 22.8 | 25.5 | 11.7 | 9.51 | 7.8 | 0.546 | 21.9 | 99.8 |
| Vaccine programme costs (discounted; \$ millions) | 124<br>(117, 136) | 470<br>(452, 495) | 648<br>(621, 687) | 173<br>(169, 178) | 127<br>(124, 131) | 24.4<br>(22.9, 26.6) | 107<br>(104, 112) | 1,680<br>(1,640, 1,720) |
| Acute healthcare costs (discounted; \$ millions) | -7.93<br>(-17.4, -3.74) | -54.6<br>(-119, -24.9) | -155<br>(-352, -60.9) | -21.4<br>(-47.5, -10.7) | -27.8<br>(-56.9, -13.5) | -3.89<br>(-8.87, -1.57) | -14.3<br>(-30.5, -6.67) | -298<br>(-534, -155) |
| Long-term healthcare costs (discounted; \$ millions) | -2.95<br>(-11.3, -0.581) | -19.8<br>(-79.2, -3.78) | -33.2<br>(-117, -7.02) | -8.13<br>(-31.1, -1.63) | -10<br>(-36.6, -2.08) | -0.871<br>(-2.97, -0.185) | -4.85<br>(-18.6, -1.01) | -86.5<br>(-252, -20.6) |
| Total incremental costs (discounted; \$ millions) | 113<br>(96.6, 127) | 394<br>(286, 446) | 456<br>(200, 581) | 143<br>(99.5, 160) | 88.9<br>(39.5, 110) | 19.6<br>(13, 23.2) | 87.9<br>(60.6, 99.7) | 1,290<br>(948, 1,490) |
| EOGBS cases (thousands) | -22.8<br>(-43.6, -11.6) | -30.6<br>(-62.4, -14.5) | -3.37<br>(-5.94, -1.57) | -8.91<br>(-16.8, -4.72) | -17.9<br>(-36, -8.6) | -0.421<br>(-0.86, -0.207) | -42.3<br>(-86.4, -20.1) | -127<br>(-248, -63.3) |
| LOGBS cases (thousands) | -11.3<br>(-33.1, -2.94) | -15.2<br>(-45.7, -3.86) | -1.99<br>(-4.2, -0.84) | -5.9<br>(-20.4, -1.94) | -12.9<br>(-33, -5.24) | -0.275<br>(-1.39, -0.0888) | -36.4<br>(-101, -14) | -87.3<br>(-209, -38.1) |
| Moderate & severe NDI cases (thousands) | -2.91<br>(-8.52, -0.933) | -3.86<br>(-11.8, -1.21) | -0.257<br>(-0.572, -0.0975) | -1.31<br>(-3.99, -0.42) | -2.63<br>(-7.58, -0.9) | -0.0541<br>(-0.223, -0.0182) | -6.66<br>(-19.4, -2.2) | -17.9<br>(-49.9, -6.38) |
| GBS deaths (thousands) | -4.22<br>(-9.6, -1.78) | -5.58<br>(-13.4, -2.32) | -0.335<br>(-0.668, -0.148) | -1.95<br>(-4.7, -0.804) | -4.76<br>(-10.8, -2.06) | -0.0836<br>(-0.336, -0.0242) | -13.6<br>(-32.2, -5.65) | -31.1<br>(-66.4, -14.4) |
| GBS stillbirths (thousands) | -6.59<br>(-23, -1.63) | -3.13<br>(-10.9, -0.794) | -0.586<br>(-1.48, -0.206) | -1.34<br>(-8.35, -0.222) | -1.33<br>(-3.12, -0.591) | -0.0646<br>(-0.336, -0.0174) | -9.31<br>(-18.5, -4.12) | -23<br>(-56.4, -10) |
| GBS associated preterm births* (thousands) | -34.7<br>(-78.5, -2.33) | -28.7<br>(-66.1, -2.05) | -33.2<br>(-72, -2.4) | -18.5<br>(-40.9, -1.37) | -23.8<br>(-52.4, -1.81) | -1.45<br>(-3.27, -0.103) | -44.4<br>(-97.4, -3.23) | -185<br>(-407, -13.5) |
| QALYs from averted GBS disease (discounted; thousands) | 150<br>(66.3, 339) | 203<br>(86.9, 485) | 12.5<br>(5.64, 24.8) | 70.2<br>(30.7, 168) | 164<br>(73, 371) | 2.96<br>(1.01, 10.6) | 431<br>(186, 1,010) | 1,060<br>(486, 2,270) |
| QALYs from averted stillbirths (discounted; thousands) | 180<br>(43.7, 627) | 87.2<br>(21.7, 301) | 16.5<br>(5.77, 41.3) | 37.3<br>(6.18, 233) | 36.9<br>(16.3, 85.9) | 1.75<br>(0.474, 9.06) | 243<br>(108, 486) | 622<br>(271, 1,550) |
| QALYs from averted preterm births* (discounted; thousands) | 63.6<br>(5.01, 162) | 53.8<br>(4.24, 143) | 62.4<br>(4.99, 160) | 34.6<br>(2.77, 88.8) | 44.2<br>(3.58, 113) | 2.77<br>(0.213, 7.23) | 77.4<br>(6.26, 198) | 338<br>(27.6, 857) |
| QALYs from averted GBS disease (undiscounted; thousands) | 358<br>(158, 807) | 513<br>(220, 1,220) | 32.7<br>(14.6, 64.6) | 177<br>(77, 422) | 408<br>(181, 923) | 6.94<br>(2.46, 24.5) | 947<br>(407, 2,230) | 2,490<br>(1,160, 5,370) |
| QALYs from averted stillbirths (undiscounted; thousands) | 429<br>(104, 1,490) | 218<br>(54.4, 752) | 42.7<br>(15, 107) | 94.1<br>(15.6, 586) | 91.4<br>(40.4, 213) | 4.19<br>(1.17, 20.9) | 532<br>(237, 1,060) | 1,460<br>(630, 3,670) |
| QALYs from averted preterm births* (undiscounted; thousands) | 152<br>(12, 387) | 135<br>(10.7, 359) | 163<br>(13, 417) | 87.8<br>(7.03, 225) | 110<br>(8.91, 282) | 7.3<br>(0.56, 19) | 171<br>(13.8, 436) | 825<br>(67.4, 2,090) |

Globally, the vaccine programme could avert an estimated 127,000 (UR: 63,300 – 248,000) EOGBS cases and 87,300 (UR: 38,100 – 209,000) LOGBS cases, thus avoiding 31,100 (UR: 14,400 – 66,400) infant deaths and 17,900 (UR: 6,380 – 49,900) cases of moderate and severe NDI. Additionally, 23,000 (UR: 10,000 – 56,400) GBS stillbirths could be prevented and, if a vaccine also proves effective against GBS associated prematurity, 185,000 (UR: 13,500 – 407,000) preterm births might be avoided. The highest burden of iGBS cases and deaths, around two-fifths of the total, is averted in Sub-Saharan Africa which accounts for about one-fifth of the women vaccinated. In contrast, only about 1% of the deaths occur in Europe and Northern America despite a tenth of vaccinated women being in this region.

Overall, iGBS cases averted through vaccination resulted in a projected gain of 2.5 million (UR: 1.2 – 5.4 million) undiscounted QALYs, and a further 1.5 million (UR: 0.6 – 3.6 million) QALYs when avoided stillbirths are included. A vaccine that prevents GBS-associated prematurity could add another 0.8 million (UR: 0.1 – 2.1 million) QALYs. The relative contribution of preventing iGBS, stillbirths and prematurity to the overall QALY gain varies by region. For example, in Sub-Saharan Africa, and Northern Africa and Western Asia preventing iGBS contributes the majority of the QALY gain, but in Europe and Northern America, and Central and Southern Asia avoided stillbirths make a larger contribution. In Europe and Northern America preventing preterm births might result in larger QALY gains than iGBS cases and stillbirths combined.

Using our base case assumptions about the vaccine characteristics the estimated global NMB of vaccination ranged from $1.1 billion (UR: $-0.2 – 3.9 billion) to $17 billion (UR: $9.1 – 31 billion) depending upon the normative assumptions (Fig. 2A). Including stillbirth QALYs increases the NMB by between $1.4 billion and $7.1 billion depending on the other normative assumptions made.

**Figure 2:**
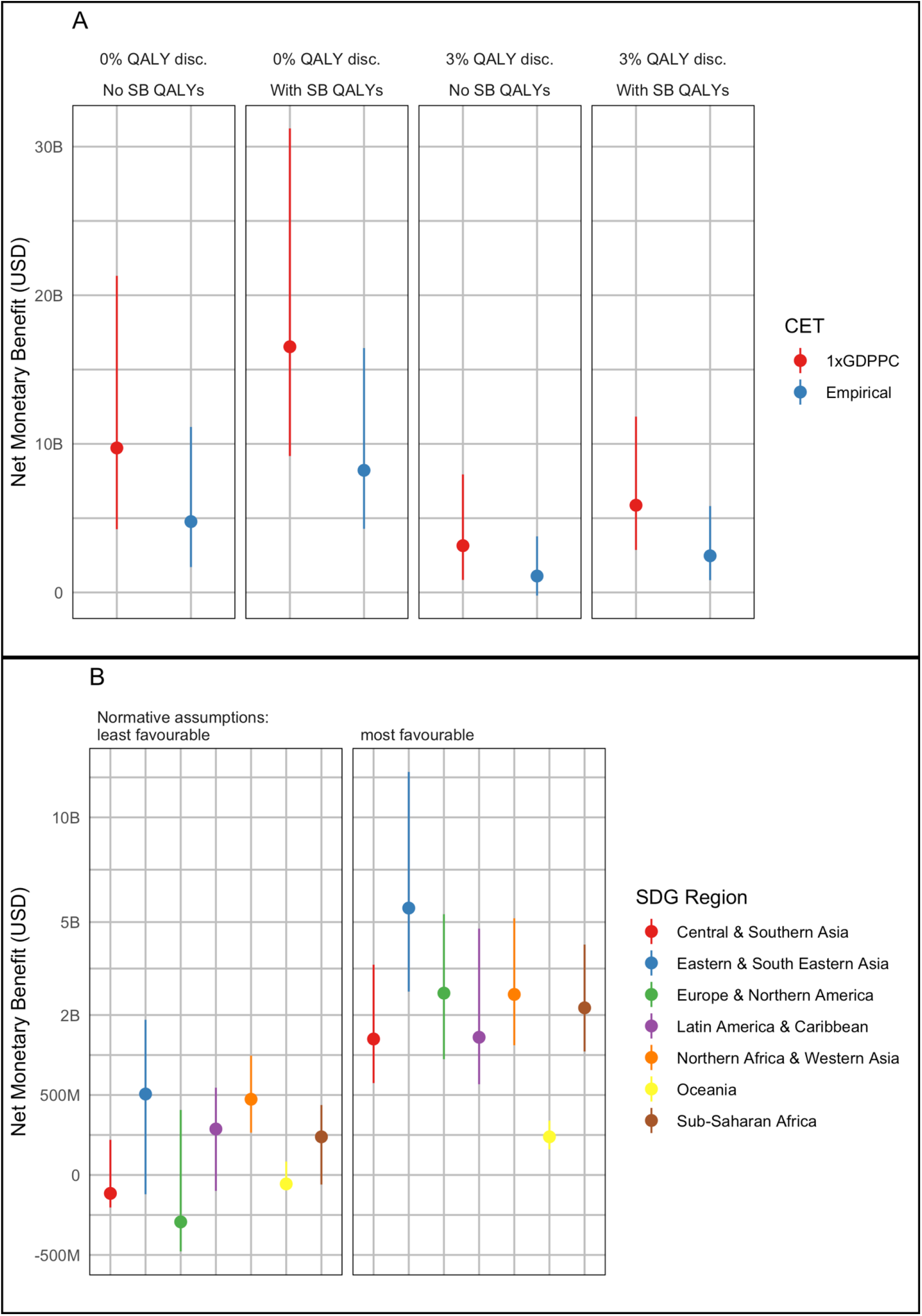
Net Monetary Benefit of GBS maternal vaccination (A) globally under different normative assumptions (see Table 1); and (B) by region for the most and least favourable normative assumptions. Least-favourable normative assumptions were the use of an empirical CET, 3% discounting of QALYs, and exclusion of stillbirth QALYs. Most-favourable assumptions were the use of 1 x GDP per capita CETs, 0% discounting of QALYs, and inclusion of stillbirth QALYs. M = Millions; B = Billions; CET = Cost-Effectiveness Threshold; GDP = Gross Domestic Product; GDPPC = GDP per capita; QALY = Quality Adjusted Life Year; SB = Stillbirth; SDG = Sustainable Development Goal.

Under the most-favourable normative assumptions vaccination had a positive NMB in all regions (Fig. 2B). However, for least-favourable assumptions the NMB was negative for Central and Southern Asia, Europe and Northern America, and Oceania. Nevertheless, if stillbirth QALYs were included, the NMB for these regions was again positive (Supplementary Fig. 1).

For the most-favourable normative assumptions vaccination is likely cost-effective in almost all countries (Fig 3.), but for least-favourable assumptions this was reduced to just over half (103/183) of countries. Notably, vaccination was less likely to be cost-effective amongst countries with lower GDP per capita within the Sub-Saharan Africa, Central & Southern Asia, and Europe & Northern America regions.

**Figure 3:**
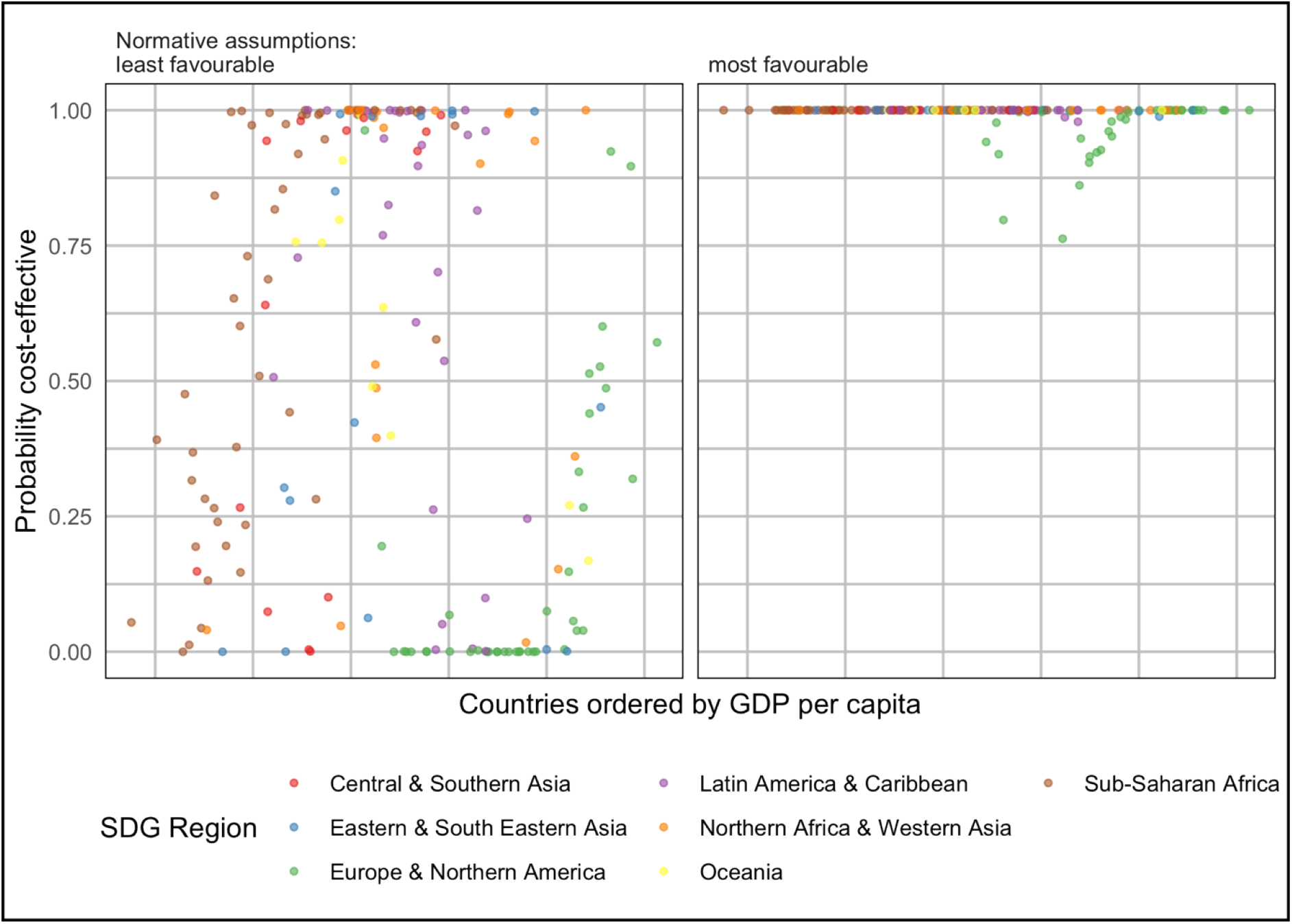
Probability that GBS maternal vaccination is cost-effective in each country under most favourable and least favourable normative assumptions (see Table 1). Least-favourable normative assumptions were the use of an empirical CET, 3% discounting of QALYs, and exclusion of stillbirth QALYs. Most-favourable assumptions were the use of 1 x GDP per capita CETs, 0% discounting of QALYs, and inclusion of stillbirth QALYs. CET = Cost-Effectiveness Threshold; GDP = Gross Domestic Product; GDPPC = GDP per capita; QALY = Quality Adjusted Life Year; SB = Stillbirth; SDG = Sustainable Development Goal.

Figure 4 shows how the global NMB of vaccination varies under different scenarios. Inclusion of mild NDI, assuming GBS births without skilled birth attendants have 90% case-fatality or increasing vaccine efficacy from 80% to 90% slightly increase the global NMB of vaccination, while assuming zero long-term costs for NDI slightly decreases the NMB. However, none of these assumptions have a dramatic effect. If vaccine efficacy is decreased to 60%, global NMB remains positive under least-favourable assumptions, but the number of countries for which vaccination is no longer cost-effective increases to 96 (Supplementary Fig. 4). A vaccine that requires two doses to achieve 80% efficacy would have a negative global NMB, and vaccination would not be cost-effective in 111 countries. However, a vaccine with protection against preterm birth substantially increases the global NMB and is especially influential in the Europe & Northern America region (see Supplementary Fig. 2).

**Figure 4:**
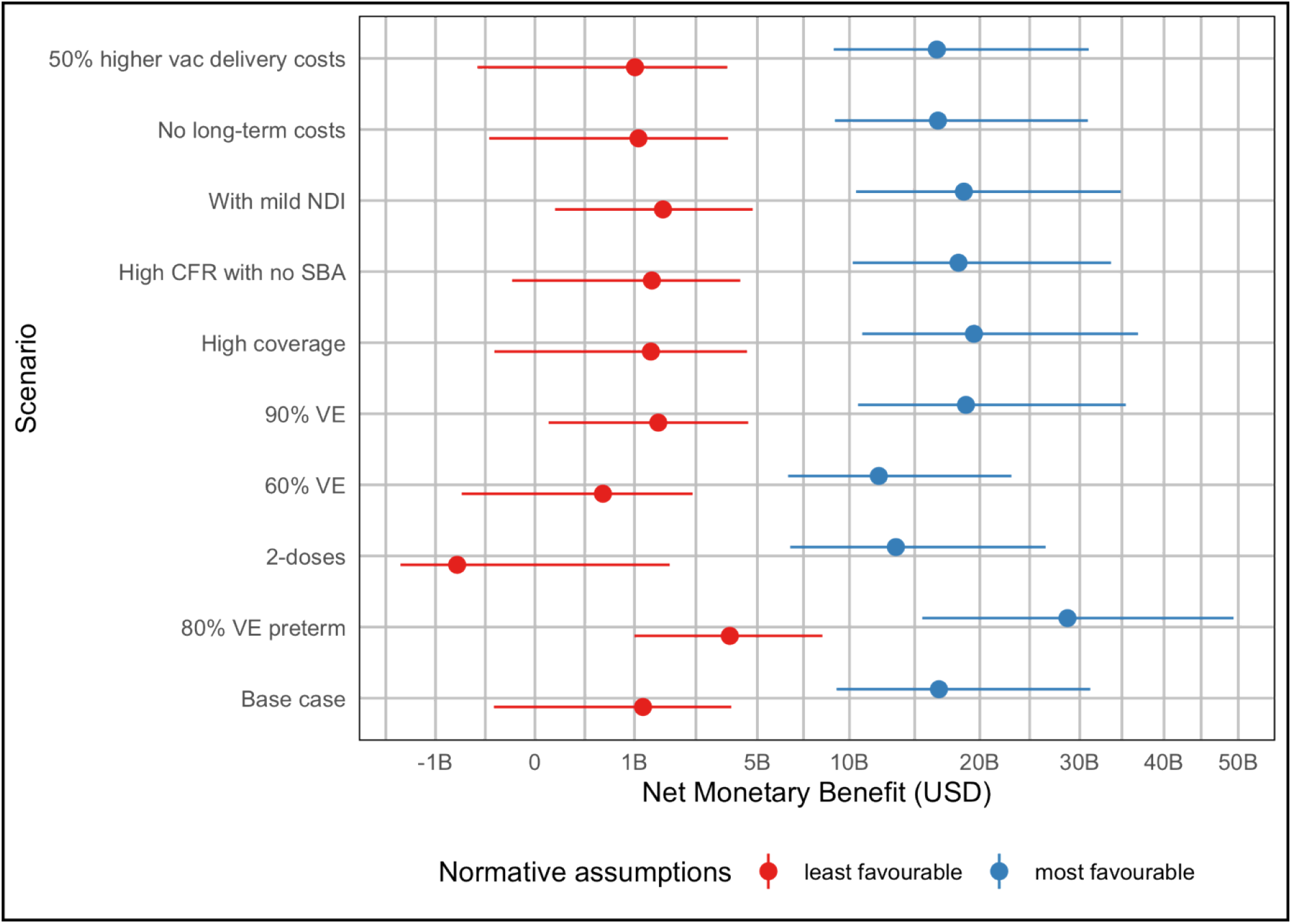
Annual global Net Monetary Benefit of GBS maternal vaccination under most favourable and least favourable normative assumptions (see Table 1) for different vaccination scenarios. Points show median estimates and lines show 95% uncertainty ranges. Least-favourable normative assumptions were the use of an empirical CET, 3% discounting of QALYs, and exclusion of stillbirth QALYs. Most-favourable assumptions were the use of 1 x GDP per capita CETs, 0% discounting of QALYs, and inclusion of stillbirth QALYs. B = Billions; CET = Cost-Effectiveness Threshold; CFR = Case Fatality Risk; GDP = Gross Domestic Product; NDI = Neurodevelopmental Impairment; SBA = Skilled Birth Attendant; QALY = Quality Adjusted Life Year; VE = Vaccine Effectiveness.

The distribution of vaccine threshold prices amongst countries within each World Bank income group are shown in Figure 5 (results by SDG region are shown in Supplementary Fig. 5, and for other vaccine scenarios in Supplementary Fig. 6). The threshold price is usually positive (i.e., there is some price at which purchasing the vaccine would be cost-effective), and generally higher in high-income and upper-middle-income countries. However, under least-favourable normative assumptions threshold price is negative in eight countries, indicating that even with a free vaccine the delivery costs outweigh the health benefits in this analysis.

**Figure 5:**
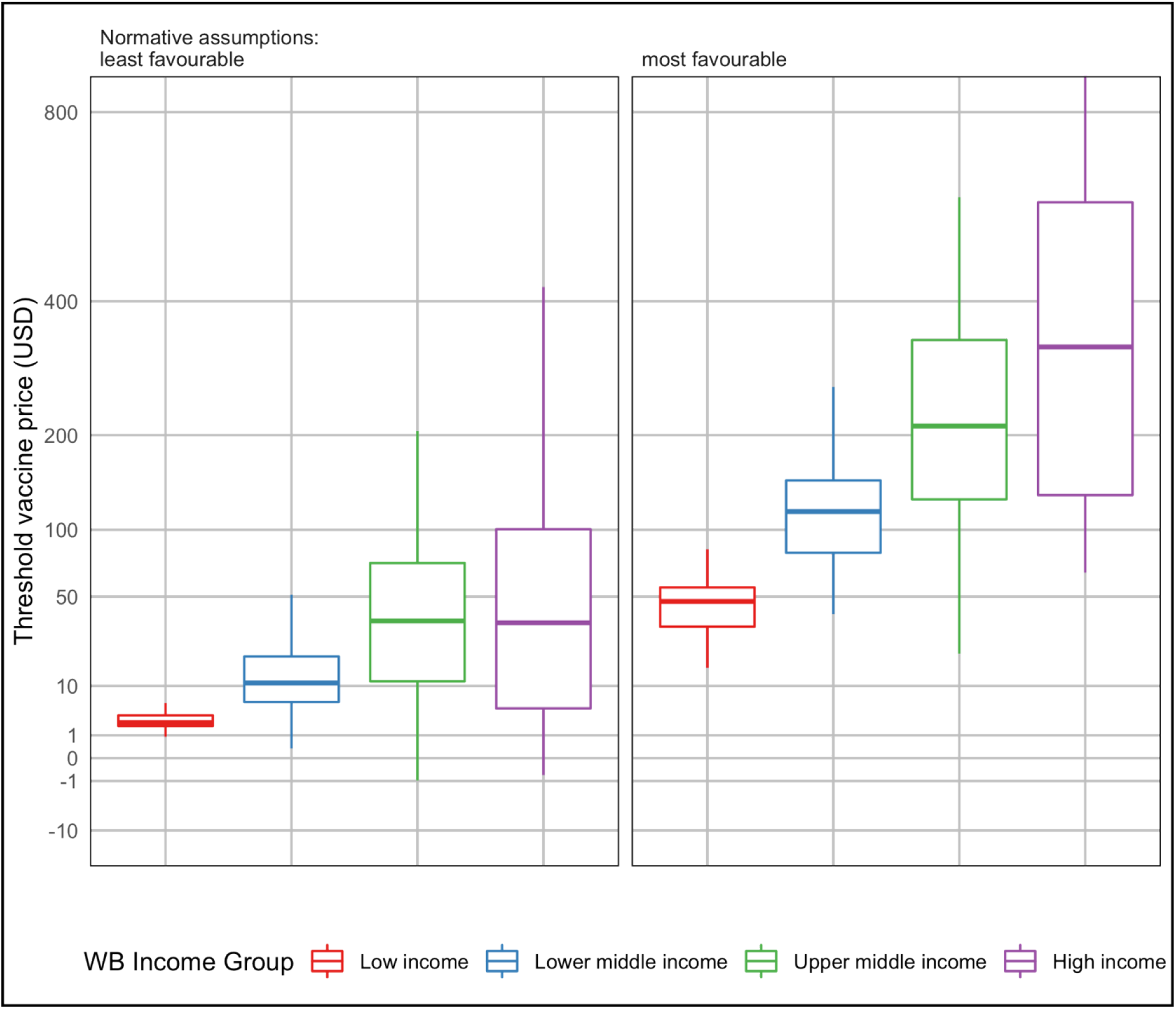
Distribution of GBS vaccine threshold prices amongst countries within each World Bank income group under most and least favourable normative assumptions (see Table 1). Threshold vaccine prices above $800 per dose are not shown. Least-favourable normative assumptions were the use of an empirical CET, 3% discounting of QALYs, and exclusion of stillbirth QALYs. Most-favourable assumptions were the use of 1 x GDP per capita CETs, 0% discounting of QALYs, and inclusion of stillbirth QALYs. CET = Cost-Effectiveness Threshold; GDP = Gross Domestic Product; QALY = Quality Adjusted Life Year.

## DISCUSSION

A high-coverage global maternal immunisation programme against GBS could avert hundreds of thousands of GBS cases, alongside tens of thousands of deaths, stillbirths, and cases of long-term disability. We estimate that such a programme may have a net cost of around $1.4 billion, with most costs occurring in Europe and Northern America. Nevertheless, it would be cost-effective in most countries under favourable assumptions, particularly if it can reduce preterm births.

Even under less favourable assumptions, a single-dose GBS vaccine could still be cost-effective due to additional factors we did not explore. In some high-income countries, GBS vaccination plus current practice may be less cost-effective compared to current practice alone because of lower GBS incidence in babies due to IAP. However, GBS vaccination might allow high-income countries to achieve additional cost savings by revising IAP algorithms for vaccinated mothers. In low- and lower-middle income countries, iGBS incidence may be higher, but so is the health opportunity cost of healthcare spending due to budget constraints leading to lower thresholds at which interventions may be considered cost-effective.

More competitive pricing may enable vaccination to be cost-effective, even under least favourable assumptions. Competitive and finely tiered vaccine prices could also be beneficial for manufacturers, with financial analyses suggesting that high global demand is needed to ensure the development costs of a GBS vaccine to be recouped.^46^ Our economic evaluation can inform both manufacturers and donors investigating the financial viability of investing in GBS vaccine development, as well as countries identifying the price they should be willing to pay for such a vaccine.

Our study is the first to estimate the value of maternal GBS vaccination across all regions and country income groups. Previous analyses have estimated cost-effectiveness in the United States,^12–14^ the Netherlands,^17^ United Kingdom,^15,16^ South Africa,^19^ The Gambia,^18^ and 37 Gavi countries in Africa.^20^ These prior estimates suggested cost-effectiveness of vaccination ranged from $320-573 per DALY averted in Gavi-eligible countries,^20^ to $3550 per DALY averted in South Africa,^19^ to over $50,000 per QALY in the United States,^12,13^ which is broadly consistent with our results. Like our analysis, Kim et al. also found that the ability to avert GBS-associated prematurity greatly improved vaccine cost-effectiveness.^19^

This was the first cost-effectiveness study to use new global estimates of the health burden due to GBS including infant morbidity and mortality, long-term neurodevelopmental impairment, stillbirth, and GBS-associated prematurity. This burden study propagated parametric uncertainty comprehensively by using a Bayesian framework to synthesise existing data sources. Posterior distributions from the study then informed a probabilistic sensitivity analysis for our cost-effectiveness model. Similarly, for cost data, parameters with multiple sources of data from previous systematic reviews were synthesised using regression models. Conversely, the main limitations of our analysis reflected parameters with limited data such as those governing health-related quality of life and long-term costs from disability, where estimates were based on only 1-2 relevant studies. Our analysis also excluded the potential impact of vaccination on maternal morbidity and the costs of GBS-related disability beyond the health-sector. However, both these factors would likely reinforce our main findings on cost-effectiveness.

A further set of uncertainties govern GBS vaccine characteristics such as efficacy, number of doses needed and impact on GBS-associated prematurity. Since there is currently no licensed vaccine, these parameters were informed by the WHO PPC which is based on expert assumptions. We therefore used scenario sensitivity analyses to identify which of these characteristics are the most important drivers of vaccine value. Further data from carefully designed vaccine trials and other field studies are needed to inform these data gaps. Vaccine value is also driven by normative health economic assumptions around discounting, cost-effectiveness thresholds and the value of preventing stillbirths, which reflect uncertainty about the values of society rather than about empirical data.

Overall, our results suggest high coverage of a competitively priced maternal GBS vaccine has the potential to save tens of thousands of lives globally and is likely to be a cost-effective investment, particularly if the vaccine can reduce GBS-associated prematurity.

## Data Availability

Code and data used in this analysis are available at https://github.com/mert0248/GBS-vax-econ-model

## Contributors’ statement

Conceptualisation – SRP, AK, JEL, and MJ; Methodology – SRP, BG, CT and MJ; Investigation – SRP; Formal Analysis – SRP; Software – SRP; Validation – SRP; Data Curation – SRP and BG; Writing – Original Draft – SRP and MJ; Writing – Review & Editing – SRP, BG, PP, JC, FS, AK, RH, CT, JEL and MJ; Visualisation – SRP; Supervision – MJ and JEL; Funding Acquisition – MJ and JEL.

## Declaration of interests

FS is employed by the UK NSC which developed the policy recommendation for maternal GBS screening.

## Acknowledgements

We would like to thank the authors of the GBS burden paper for sharing data on the posterior estimates of parameters in the burden model. We thank the GBS Full Value of Vaccine Assessment project Scientific Advisory Group for helpful discussion. We also thank Clint Pecenka and Ranju Baral for sharing estimates of antenatal care coverage by gestational age. This work was supported by a grant (INV-009018) to the London School of Hygiene & Tropical Medicine (PI Joy Lawn) from the Bill & Melinda Gates Foundation. RH is member of the WHO. The views expressed in this article are those of the authors and do not necessarily represent the decisions, official policy or opinions of the WHO.

## Data sharing statement

Code and data used in this analysis are available at https://github.com/mert0248/GBS-vax-econ-model

## Notes

### Competing Interest Statement

I have read the journal's policy and the authors of this manuscript have the following competing interests: FS is employed by the UK NSC which developed the policy recommendation for maternal GBS screening. RH is member of the World Health Organisation. The views expressed in this article are those of the authors and do not necessarily represent the decisions, official policy or opinions of the WHO.

### Funding Statement

SRP, BPG, PP, JC, FS, AK, JEL and MJ were supported by funding from a grant (INV-009018) to the London School of Hygiene & Tropical Medicine (PI Joy Lawn) from the Bill & Melinda Gates Foundation. The funders had no role in study design, data collection and analysis, decision to publish, or preparation of the manuscript.

